# Meta-analysis Of Transcriptomic Data Reveals Pathophysiological Modules Involved With Atrial Fibrillation

**DOI:** 10.1101/2020.05.13.20100867

**Authors:** Rodrigo Haas Bueno, Mariana Recamonde-Mendoza

## Abstract

Atrial fibrillation (AF) is a complex disease and affects millions of people around the world. The biological mechanisms that are involved with AF are complex and still need to be fully elucidated. Therefore, we performed a meta-analysis of transcriptome data related to AF to explore these mechanisms aiming at more sensitive and reliable results. Public transcriptomic datasets were downloaded, analyzed for quality control, and individually pre-processed. Differential expression analysis was carried out for each individual dataset, and the results were meta-analytically aggregated using the r-th ordered p-value method. We analyzed the final list of differentially expressed genes through network analysis, namely topological and modularity analysis, and functional enrichment analysis. The meta-analysis of transcriptomes resulted in 589 differentially expressed genes, whose protein-protein interaction network presented 11 hubs-bottlenecks and four main identified functional modules. These modules were enriched for, respectively, 23, 54, 33, and 53 biological pathways involved with the pathophysiology of AF, especially with the disease’s structural and electrical remodeling processes. Stress of the endoplasmic reticulum, protein catabolism, oxidative stress, and inflammation are some of the enriched processes. Among hubs-bottlenecks genes, which are highly connected and probably have a key role in regulating these processes, we found *HSPA5, ANK2, CTNNB1*, and *VWF*. Further experimental investigation of our findings may shed light on the pathophysiology of the disease and contribute to the identification of new therapeutic targets and treatments.

## INTRODUCTION

Atrial fibrillation (AF) is the most common type of cardiac arrhythmia and affects the life quality of millions of people around the world. With the global ageing population phenomenon, a considerable increase in the number of AF diagnoses is expected, since its prevalence and incidence rises with age (Heeringa et al. 2006). AF is related to an increased risk of thromboembolic events (strokes and ischemic heart disease), and has a high prevalence in patients with other cardiovascular or metabolic diseases such as hypertension, heart failure, or diabetes mellitus (Ball et al. 2013). AF is considered a chronic and progressive disease, and its pathophysiology involves structural and electrophysiological changes in the atrial tissue that, collectively, promote the formation and propagation of abnormal impulses through the atria (Nattel and Harada 2014).

Despite advances in revealing the mechanisms behind AF, there are still numerous gaps to be filled (Heijman et al. 2014)(Sühling et al. 2018). In this context, for more than a decade, analysis of transcriptomic data has proved to be useful in the identification of genes and biological mechanisms involved in complex diseases such as cancer (Rhodes and Chinnaiyan 2005), cardiovascular (Nanni et al. 2006), and neurological diseases (Carpanini et al. 2017), with a wide availability of microarray datasets deposited in public databases. Several studies addressed the identification of gene expression changes at the transcriptional level in response to AF, highlighting putative biomarkers and molecular mechanisms and targets that mediate AF-dependent remodeling processes (Deshmukh et al. 2015; Kim et al. 2003; Sühling et al. 2018).

However, analysis of microarray data usually suffers from low reproducibility of results and with low statistical power due to small sample size in proportion to the thousands of measured probes (Walsh et al. 2015). In addition, the complexity of the molecular mechanisms underlying the pathophysiology of AF and the large heterogeneity of AF patients adds an extra layer of complexity to the extraction of precise hypotheses about AF targets and mechanisms. Therefore, combining the results of different studies using meta-analysis methods appears as an alternative to mitigate these methodological limitations inherent to this type of data (Grills et al. 2011; Ghosh et al. 2003), and overcome the challenges imposed by disease heterogeneity.

In this study, we performed a meta-analysis of transcriptomic data involving tissue and blood samples of patients affected by AF to better understand its molecular mechanisms. Through a bioinformatics approach, we investigated potential interactions and biological processes associated with the activity of the genes returned by our meta-analysis. The biological plausibility of observed associations was evidenced by literature-based validation, with our findings corroborating several recent works that highlight key elements and pathways subjacent to the onset and development of AF. Experimental validation of our insights can uncover new pathophysiological mechanisms of this disease, as well novel candidates for therapeutic targets.

## METHODS

In this meta-analytical study, the following steps were followed: 1) Manual search of public transcriptomic data; 2) Pre-processing of transcriptomic data and differential expression analysis; 3) Meta-analysis of transcriptomic data; 4) Analysis of biological networks; 5) Interpretation of results. Steps 2 and 3 were implemented in the R environment (version 3.6.0 for Windows) (Ripley 2001). Step 4 was implemented in Cytoscape software (version 3.7.1) (Shannon et al. 2003), using specific apps available for this software. For step 5, Cytoscape (version 3.7.1), as well as public domain web tools well consolidated for the respective analyses were used.

### Manual search of public data

Search for transcriptomic data was carried out in the public Gene Expression Omnibus (GEO) database (Edgar, Domrachev, and Lash 2002). The chosen search keywords included the terms “atrial fibrillation”, “arrhythmia”, “heart”, “cardiac”, “disease”, “gene expression profiling”, and “transcriptome”. Data selection was made according to the following criteria: a) transcriptomic data generated with microarray techniques; b) studies performed on human samples; c) studies with groups and samples clearly described and with at least three samples per group; d) availability of raw data; e) data published after 2001, giving priority to those published after 2007 due to the better precision of more recent technologies. The search was conducted in April 2019.

### Pre-processing of transcriptomic data

Quality evaluation of raw data is important for subsequent analysis of microarray data, since it increases the detection capacity of differentially expressed genes (Edgar, Domrachev, and Lash 2002; Kauffmann and Huber 2010) as well as the efficacy of meta-analysis (Ramasamy et al. 2008). Quality assessment process was run on a platform-dependent basis, as follows:

- Affymetrix platform: for affymetrix chips, arrayQualityMetrics package for R was used (Kauffmann, Gentleman, and Huber 2009). This package computes, from raw data, six metrics commonly used to assess the quality of raw microarray data: distances between arrays, principal component analysis (PCA), boxplots, density plots, standard deviation versus rank of the mean, and MA-plots. In our study, samples considered outliers by the software in at least two metrics were excluded.
- Agilent Platform: for Agilent chips, quality assessment was based on the evaluation of MA-plots, background intensities boxplots and PCA, as recommended by Limma user’s guide (Ritchie et al. 2015). Extremely deviant samples in any of the metrics were excluded.

Following quality evaluation, normalization aims to adjust the effects on data from technical and environmental variations such as erroneous hybridization or labeling reactions; physical or structural problems; problems with reagents or laboratory conditions and, finally, with the scanning of hybridized chips. These corrections allow samples from the same study to be comparable to each other. Similarly to the previous step, the normalization process was applied according to the study’s platform:

- Affymetrix platform: raw data from GEO database were imported into R and processed with the Robust Multi-array Average (RMA) algorithm (Irizarry et al. 2003) through the oligo R package (Carvalho and Irizarry 2010). This algorithm performs different pre-processing steps: background correction, data transformation using a base 2 logarithm, normalization using the quantile method and summarization of the probesets in a single expression value for each gene. Probe to gene symbol annotation was done through specific annotation packages for each chip model available in the Bioconductor repository. Probes with annotations for more than one gene or that did not have annotations were excluded from the analysis. In order to model and to correct possible batch effects (source of variation mainly due to technical heterogeneity – different experimentation times, different reagent lots, etc.), the surrogate variable analysis (SVA) correction algorithm was applied to data (Carvalho and Irizarry 2010; Leek and Storey 2007).
- Agilent Platform: raw data from GEO database were imported into R and the following pre-processing steps were performed using the limma package: background correction (normexp algorithm, mle method, 50 offset (Ritchie et al. 2007; Silver, Ritchie, and Smyth 2009) and between-array normalization using the quantile method (Yang and Thorne 2003). Probe to gene symbol annotation was made from tables provided by the company responsible for the platform. Probes with annotations for more than one gene or without annotations were excluded from the analysis. For batch effects modeling, the SVA algorithm was also used.

### Differential expression analysis

To estimate the p-values from the hypothesis tests about the differential expression of each gene in each of the datasets, which are the inputs for the meta-analysis method, the well-established limma package protocol for differential expression analysis was used. Briefly, a linear model from the expression data for each gene is created and used to perform a “moderated” t-test (Smyth 2004), which stabilizes and increases statistical power of the analysis of experiments with small sample sizes.

### Meta-analysis

Meta-analysis of transcriptomic data was performed using the MetaDE R package (X. Wang et al. 2012). In this study, the meta-analysis was restricted for the genes commonly found in at least 80% of individual datasets, using the p-values ?calculated for each gene in each dataset, and their corresponding effect direction obtained from estimated effect size as described in (Marot et al. 2009). There are several statistical techniques that can integrate microarray data. They differ in the statistical parameters they use and in terms of how statistically conservative their results are (Chang et al. 2013). In general, the choice of method is based on researchers’ objectives and hypotheses, and on data heterogeneity (Ramasamy et al. 2008).

The exploratory objective of this study would be limited by very conservative methods; however, the presence of datasets of different microarray platforms, as well as different types of biological samples add considerable heterogeneity to the data. Our choice, therefore, was to use the r-th ordered p-value (roP) method, which is considered robust, consistent, and only moderately conservative by tests carried out in review of meta-analysis methods (Chang et al. 2013). The r-th parameter refers, in a simplified way, to the proportion of datasets in which a gene must have been considered differentially expressed so that this method of meta-analysis considers it as such (Song and Tseng 2014). In this study, a rth parameter equal to 0.5 was applied, meaning that at least 50% of datasets containing data on a given gene must consider it statistically differentially expressed so that the meta-analytical final result of the gene is considered differentially expressed (Song and Tseng 2014). The final p-values obtained for each gene were corrected using the false discovery rate (FDR) method Benjamini and Hochberg 1995). Only the genes considered differentially expressed with a 95% confidence level (FDR <0.05) by the rOP method and presenting a divergent effect direction in a maximum of 30% of datasets that contained information about these genes were considered for further analysis (Figure 1).

### Network analysis

Based on the list of genes with similar effect directions and considered differentially expressed by the meta-analysis, a search with a high reliability parameter (0.7) was conducted in STRING database (Szklarczyk et al. 2015) to generate a protein-protein interaction (PPI) network among genes potentially associated to AF. The basic PPI network was imported into Cytoscape application (Shannon et al. 2003), to which additional information regarding the regulation (up or down) of each gene was manually added. To evaluate the network’s topological parameters, Cytoscape’s NetworkAnalyzer (Assenov et al. 2008) plugin was used. Two measures of centrality were evaluated to determine the central nodes of the network: degree and betweenness. While degree quantifies the number of connections from a node, betweenness reflects the number of minimum paths between two nodes in the network that pass through a given node. Finally, clustering analysis to identify highly connected nodes within the network was performed using the MCODE application (Bader and Hogue 2003) for Cytoscape. This algorithm identifies densely connected regions in the target network that are candidates for representing functional interaction complexes.

### GO functional enrichment analysis

Gene ontology (GO) functional enrichment analysis (Ashburner et al. 2000; The Gene Ontology Consortium 2019) was performed on genes composing the main clusters identified by MCODE, searching for over-represented ‘biological process’ (BP) terms in each of these clusters. This analysis was conducted with the R clusterProfiler package (Yu et al. 2012), using the available over-representation statistical test.

## RESULTS

### Datasets included in the meta-analysis

According to inclusion criteria, nine datasets were selected for this meta-analysis. Six were generated with Affymetrix chips and three with Agilent chips. The datasets contained a total of 79 samples from patients with atrial fibrillation and 75 samples from patients with sinus rhythm (controls). Atrial tissue samples were used in eight datasets, while only one study used blood samples. Relevant information about datasets selected for this meta-analysis is summarized in Table 1, in which the number of samples includes only samples that passed raw data quality control. Raw data of selected datasets were downloaded and imported into the R platform for further analysis.

**Table 1.**
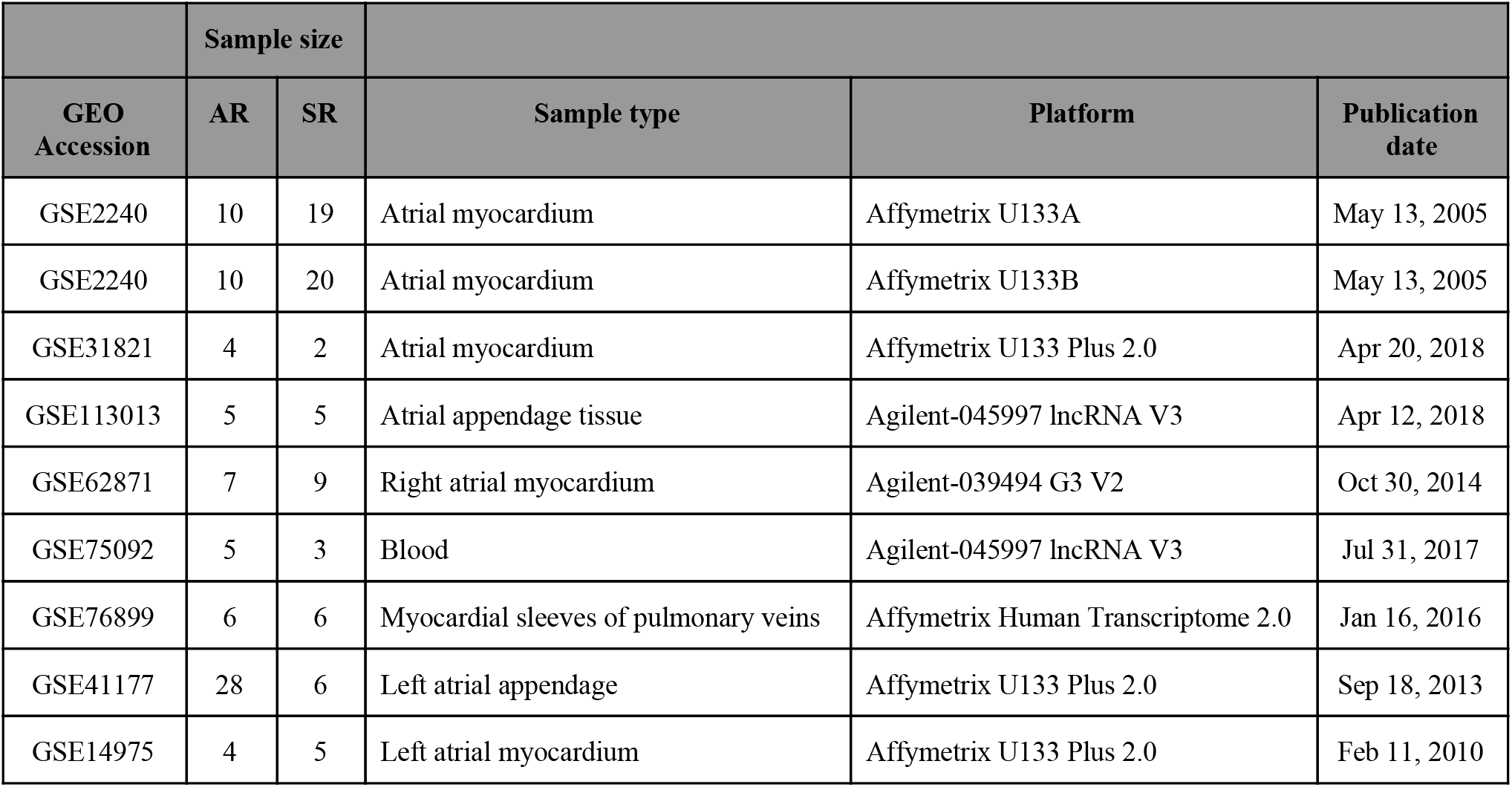
Characteristics of datasets selected for meta-analysis. AF: Atrial Fibrillation. SR: Sinus rhythm GSE2240

### Quality control of raw data

Applying raw data quality control analysis as described in the methods section, doubtful quality samples were removed from subsequent analyses. In this step, the number of samples removed by dataset were: six samples from GSE2240 (GPL96), five samples from GSE2240 (GPL97), one sample from GSE75092, four samples from GSE41177, and one sample from GSE14975.

### Individual dataset analysis and meta-analysis

Preprocessing of raw data and differential expression analysis resulted in nine lists (one list for each dataset) of genes with their respective p-values (i.e., hypothesis test about the differential expression of each gene) and direction effects. As different platforms differ in the number of transcripts measured, we carried out the meta-analysis only on data of 14582 genes represented in at least 80% of selected datasets.

In our meta-analysis, 2518 genes were considered differentially expressed in the final result using the roP method with r-th parameter equal to 0.5. However, many genes showed conflicting effect directions, i.e., up or downregulation, between datasets. In this context, 589 genes (356 upregulated and 233 downregulated, see Table S1), whose effect direction disagreed in a maximum of 30% of the datasets covering their expression remained for further analysis (Figure 1).

**Fig. 1.**
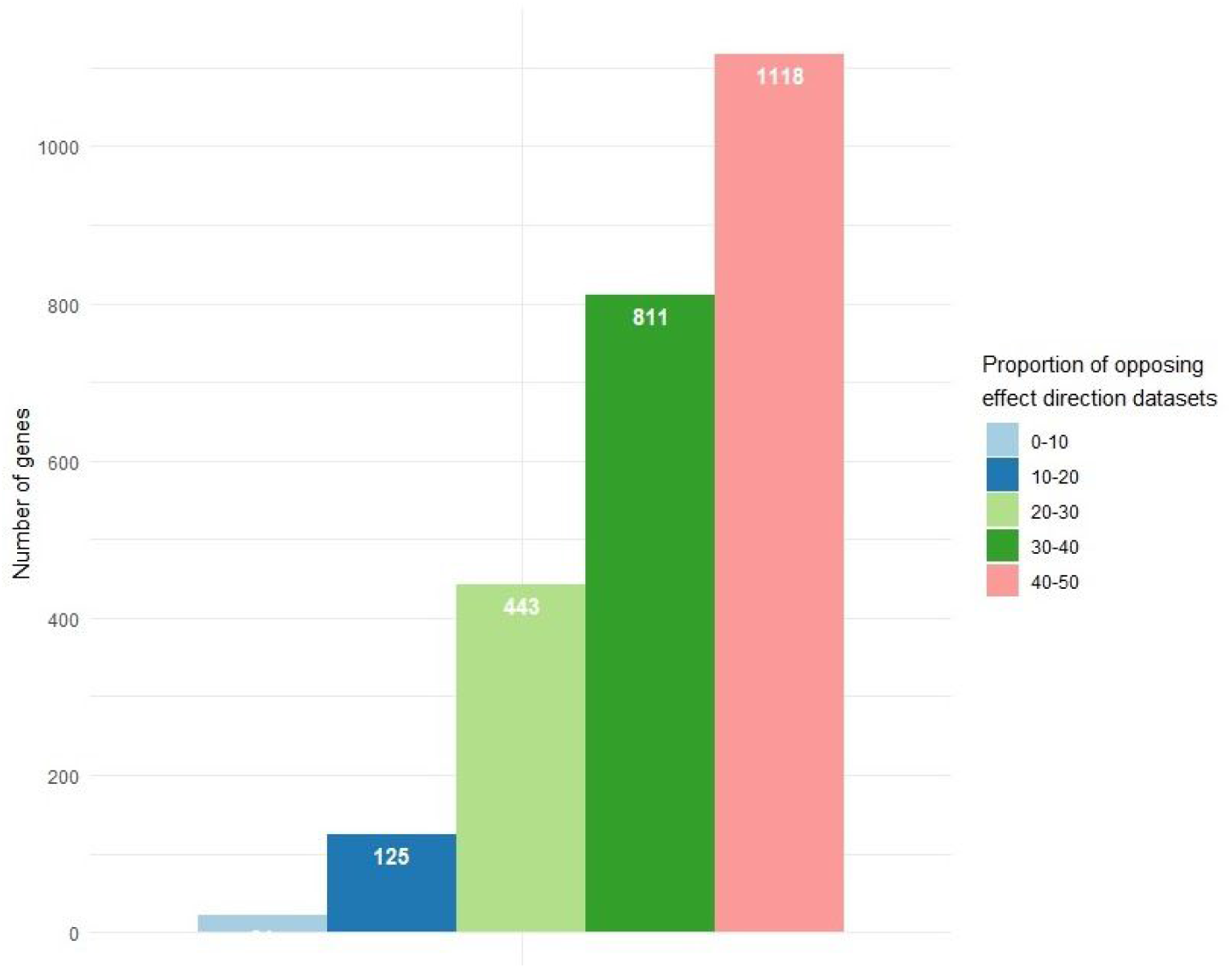
Proportion (%) of datasets with divergent effect directions of differentially expressed genes identified in our meta-analysis. Only genes with opposite effect directions in a maximum of 30% of datasets containing information about them (i.e., genes included in the light blue, blue, and light green bars) were kept, totaling 589 genes.

### Network analysis

The PPI network imported from STRING had 527 nodes and 1750 edges. Nodes represent the proteins encoded by differentially expressed genes; nodes not connected to the network were excluded. Topological analysis of degree distribution revealed a good fit (r-squared = 0.875) to the exponential model, featuring a scale-free network, like most biological networks (Barabási and Oltvai 2004). The intersection between the 15 nodes with the highest degree and the 15 nodes with the highest betweenness resulted in 11 hubs-bottlenecks nodes, which are key elements in the functional analysis of a network, since they tend to be functionally important proteins and evolutionarily conserved in the represented processes in the biological network (Park and Kim 2009). Of these, nine represent upregulated genes and 2 represent downregulated genes in AF in relation to controls (Table 2).

**Table 2.** Hubs-bottlenecks identified in the main PPI network generated from differentially expressed genes. Due to their topological properties, they represent interesting candidates to play an important role in the pathophysiology of AF.

| Network Node | Betweenness-Centrality | Degree | Regulation |
| --- | --- | --- | --- |
| SRSF1 | 0.0509 | 23 | Upregulated |
| NPM1 | 0.0572 | 25 | Upregulated |
| CLTC | 0.0353 | 24 | Upregulated |
| CCNB1 | 0.0481 | 30 | Upregulated |
| BDNF | 0.0459 | 28 | Upregulated |
| ANK2 | 0.0268 | 22 | Downregulated |
| GART | 0.0635 | 29 | Upregulated |
| VWF | 0.0294 | 22 | Upregulated |
| HSPA5 | 0.0975 | 38 | Downregulated |
| NRAS | 0.0464 | 34 | Upregulated |
| CTNNB1 | 0.0982 | 45 | Upregulated |

**Figure 2.**
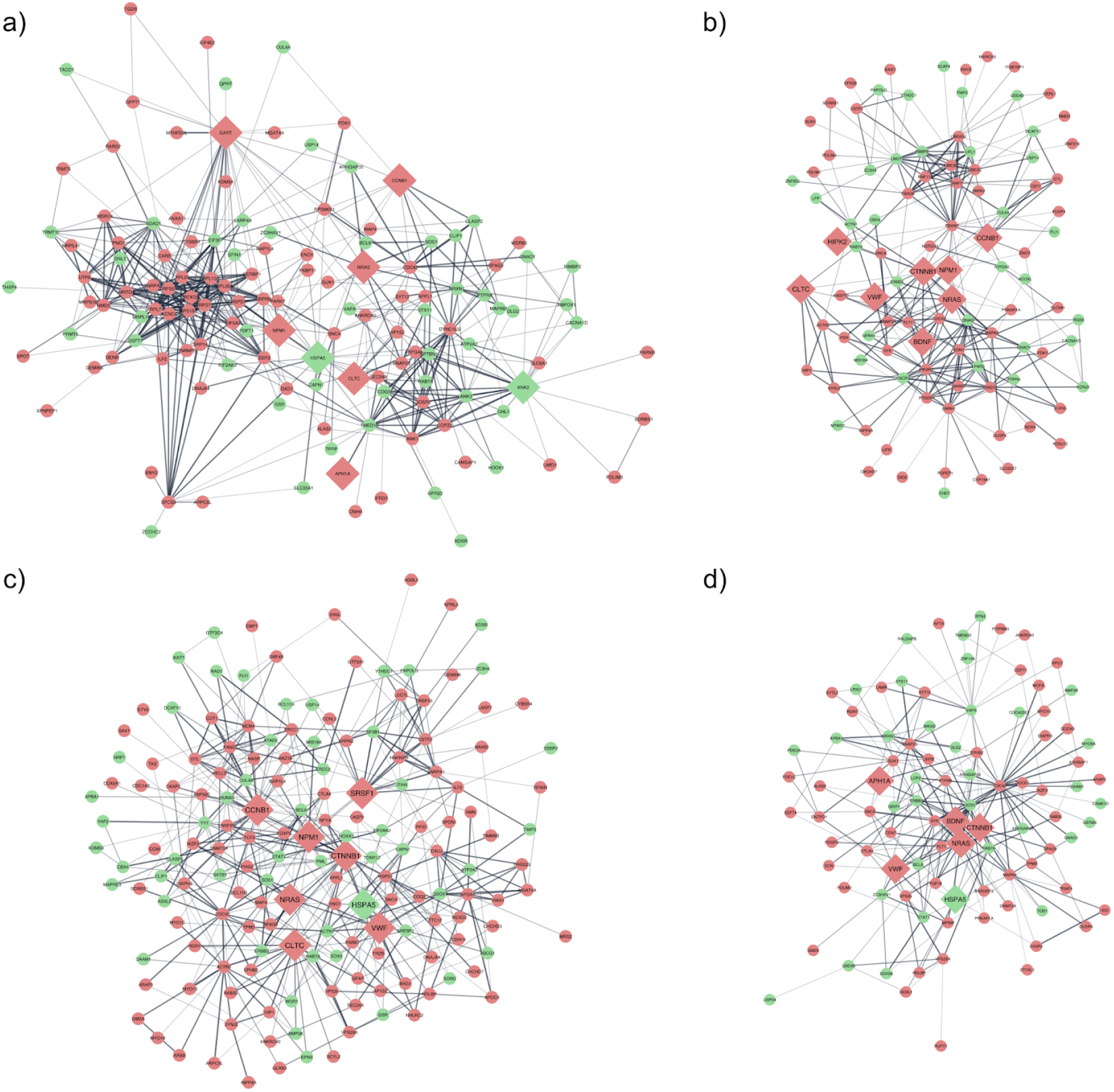
The four modules with the highest score identified by MCODE in the PPI network. Green nodes denote down-regulated genes in AF, whereas red nodes represent up-regulated genes. Diamond nodes indicate hub-bottlenecks identified in the network structural analysis.

Another important characteristic of biological networks is their functional modules, whose identification can reveal coordinated biological processes, since proteins that occur in the same cluster tend to enrich for common biological functions (Charitou, Bryan, and Lynn 2016). From the main network, 13 clusters were detected using the MCODE program. The four modules with the highest score, cluster 1 (7.74), cluster 2 (5.96), cluster 3 (5.72), and cluster 4 (4.90) contain, respectively, 132, 98, 162, and 94 genes, each of which may be interpreted as functional modules.

### Functional enrichment of clusters

To identify biological pathways enriched in the modules of our network, the over-representation statistical test was applied to the constituents of each module using GO – Biological Process database. Modules 1, 2, 3, and 4 enriched, respectively, for 2832, 3367, 3657, and 2796 statistically significant (FDR < 0.05) biological pathways. To simplify the interpretation of these results, an algorithm provided by the R ClusterProfiler package(Yu et al. 2012) for summarizing the redundant terms of the biological pathways’ names was used. Thus, the final number of enriched and summarized pathways for modules 1, 2, 3, and 4 were, respectively, 23, 54, 33, and 53. To eliminate the presence of enriched pathways with few genes in relation to the total number of component genes of the respective modules (gene ratio), pathways of each module with a gene ratio lower than the 30th percentile of the gene distribution of such module were excluded. The final list of enriched pathways for each module, their respective gene ratios, and adjusted p-values ?are shown in Figure 3 (see Table S2 for further information, including clusters’ deregulated transcripts). We observed that enriched terms have a small overlap between different clusters, whereas for a given cluster we noted a relatively high intersection between genes involved in its enriched pathways (Figure S1). This reinforces the idea of functional modularity in the network, since there is a certain degree of functional independence between clusters, but not intra-cluster. (Figure S1).

**Figure 3.**
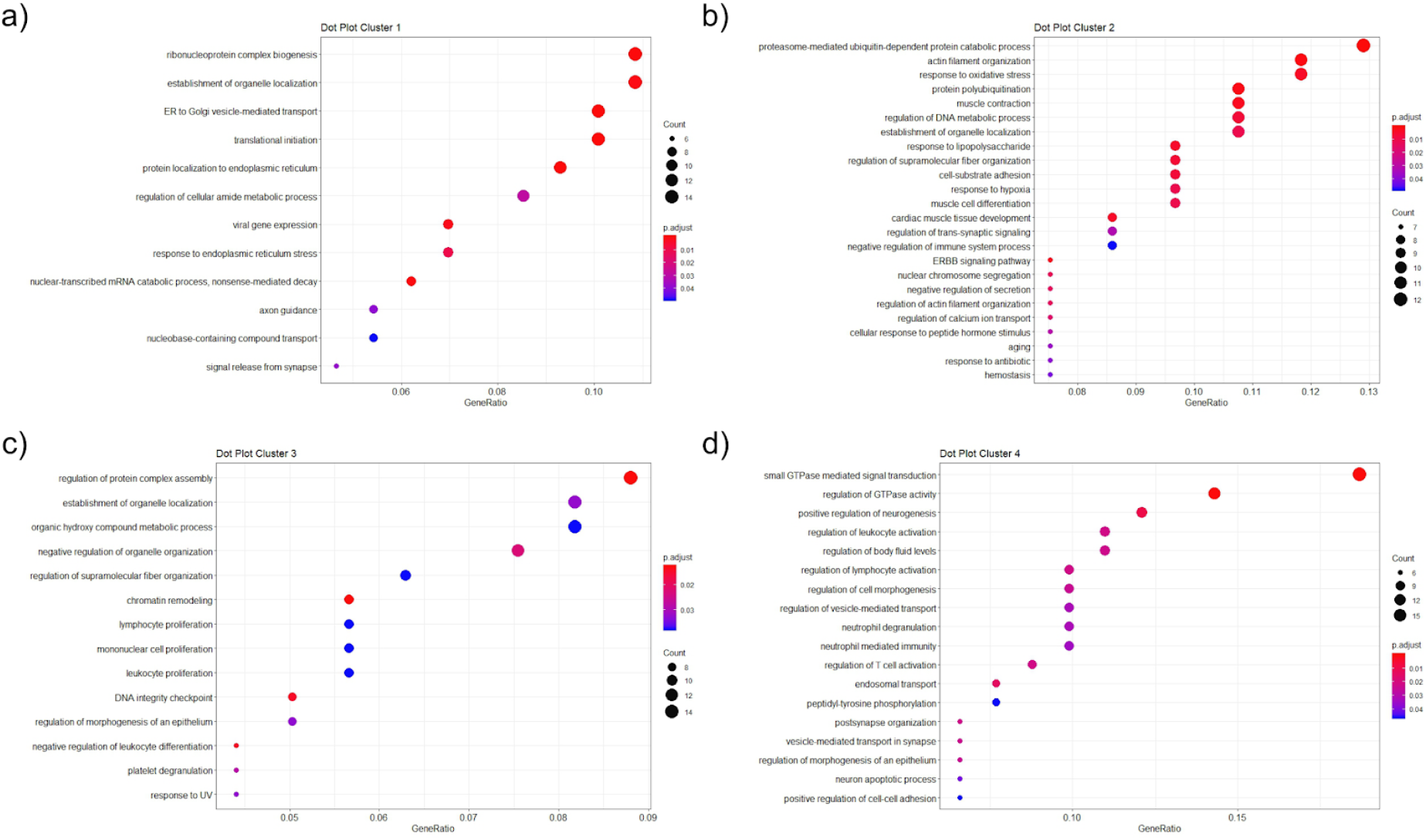
Biological pathways enriched for differentially expressed genes in each of the functional modules identified by cluster analysis, namely, a) Cluster 1, b) Cluster 2, c) Cluster 3, and d) Cluster 4. Circle size represents the number of genes enriching for a given biological pathway and the color of the circle represents statistical significance, with red denoting higher statistical significance. GeneRatio is the ratio between the number of genes that have been enriched for a given pathway and the total number of genes that belong to the respective module.

## DISCUSSION

AF is a complex and progressive disease that involves multiple pathways of electrical and structural remodeling of the atrial tissue, which culminate in the emergence of ectopic activity and dysfunctional conduction of the cardiac impulse in the atria. To advance the knowledge about the mechanisms that permeate it, this study used nine public transcriptomic data and performed a meta-analysis of differential expression, obtaining a list of 589 genes differentially expressed in a consistent manner between the individual data sets. The interaction network between the protein products of these genes, in turn, revealed functional modules whose enriched biological processes represent important and interrelated processes in tissue and in the pathophysiological context of AF.

The enriched biological processes for the protein products of genes belonging to the different modules identified in the network are shown in Figure 4. As expected, the modules have little overlap of biological pathways, indicating a modular and functional behavior of the pathophysiological processes involved with AF, but still presenting some interface between the different modules that may suggest communicating links between them. Module 1 represents, among others, biological pathways related to protein synthesis – initiation of translation and biogenesis of ribosomes – and to the physiology of endoplasmic reticulum (ER) – protein localization, vesicle transport to the Golgi complex and stress response of ER. The cellular functions of protein synthesis and folding and calcium homeostasis performed by ER are inseparable from the pathological processes of structural remodeling of cardiomyocytes present in AF (Wiersma et al. 2017). A series of harmful stimuli related to AF such as calcium depletion, hypoxia, oxidative stress, inflammation, exacerbation of protein synthesis and myolysis, as well as aging, can trigger ER stress and subsequent cellular responses, culminating, for example, in the expression of heat-shock proteins (HSPs) genes (Nattel and Harada 2014; Amen et al. 2019). These proteins attenuate the deregulation process of the dynamics of synthesis/degradation and of the function of structural and contractile proteins of cardiomyocytes in AF (Allessie et al. 2010), facilitating folding/refolding of proteins and degradation of protein aggregates that can lead to cardiomyocyte death (Amen et al. 2019). HSPA1A and members of the HSPB family are examples of chaperones whose presence protects against structural remodeling of cardiomyocytes and for which the pharmacological induction of expression has been evaluated (Hoogstra-Berends et al. 2012). The topological analysis of our main network revealed the chaperone BiP (binding immunoglobulin protein), encoded by the *HSPA5* gene, as a hub-bottleneck, probably playing a key role in the pathophysiological dynamics of AF. In addition to the classic functions of a chaperone, BiP plays a signaling role in protective cellular responses against ER stress, as well as a regulatory role for intracellular calcium dynamics (J. Wang et al. 2017). Our study identified *HSPA5* gene transcript as consistently downregulated. Overexpression of this protein was able to attenuate stress-induced apoptosis of ER, while its absence causes leakage of calcium from ER through Sec61-channel complex (Fu et al. 2008; Schäuble et al. 2012). Thus, the low expression of the *HSPA5* gene transcript may be failing to play a protective role against structural remodeling and apoptosis in cardiomyocytes subjected to stress in AF, as well as contributing to ectopic depolarization of atrial tissue via induced delayed afterdepolarizations (DADs) by leakage of calcium from sarcoplasmic reticulum (SR). BiP’s expression modulation, therefore, may prove to be a promising mechanism in interrupting structural and electrical remodeling of atrial tissue, arresting the progress of AF.

**Figure 4.**
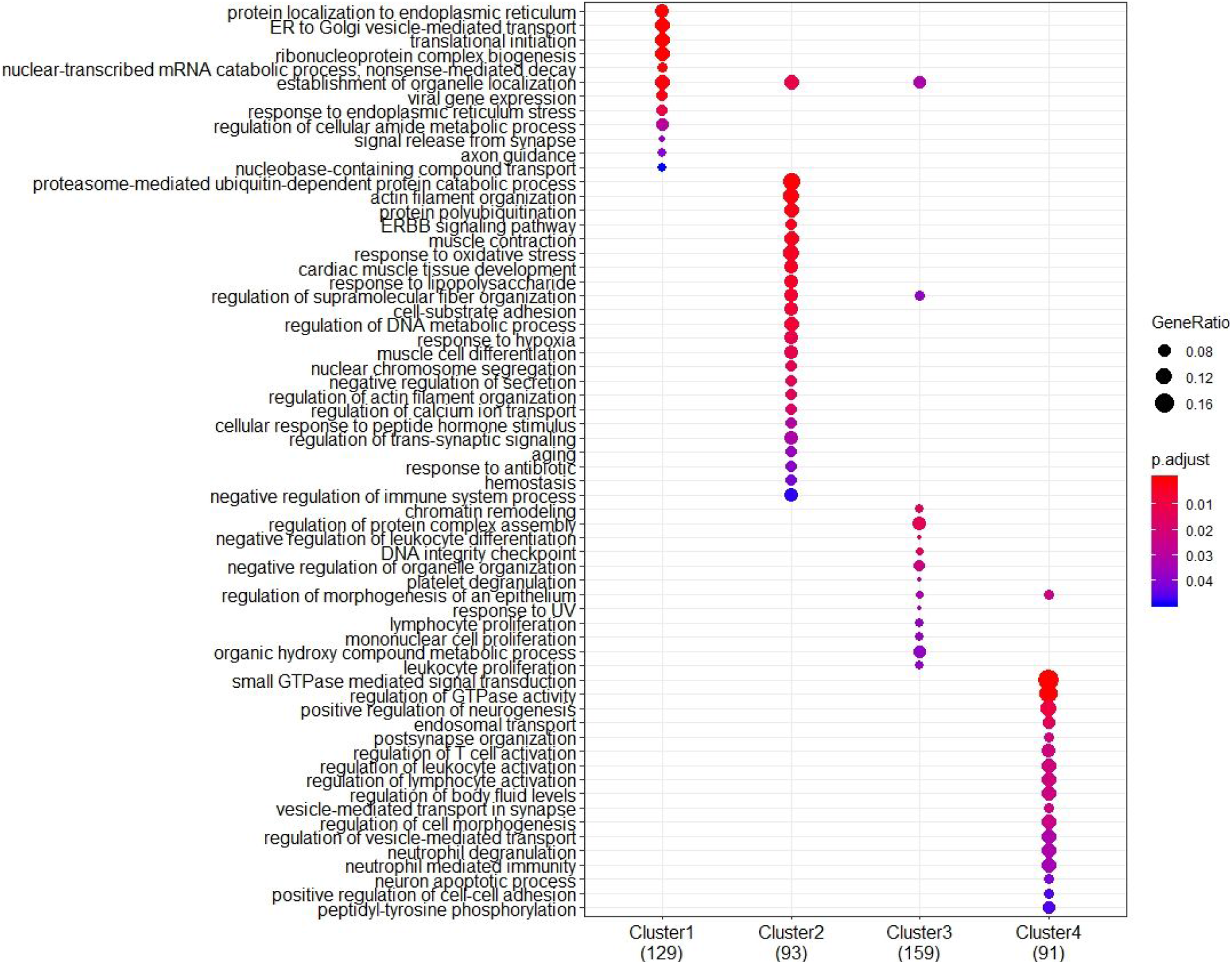
Comparison between enriched pathways for each of the main modules identified. Interestingly, enriched pathways have little overlap among modules, indicating that they are functionally distinct.

Another downregulated gene related to the enriched pathways for ER physiology/protein localization and considered a hub-bottleneck by the topological analysis of the main network of deregulated genes identified by our meta-analysis is *ANK2*. This gene codes for a series of isoforms of ankirin-B protein. In the cellular context, these proteins are adapter links between integral membrane proteins and cell cytoskeleton, organizing structural, electrogenic and signaling protein networks. The loss of function of *ANK2* gene has already been associated with several cardiac pathologies, including abnormalities in cardiac impulse conduction and arrhythmias, including AF (Cunha and Mohler 2009). In cardiomyocytes, ankirin-B appears to organize a complex involving the *α*2 isoform of the Na^+^/K^+^ pump (NKA) and the Na^+^/Ca^2+^ exchanger (NCX) in a microenvironment in which NCX activity is regulated by NKA. In this context, the loss of function or low expression of ankirin-B disturbs the regulation and localization in the membrane of these exchangers, increasing the rates of Ca^2+^waves associated with SR leakage via ryanodine receptor (RyR) (Skogestad et al. 2020), an important mechanism that causes DADs (Shiferaw, Aistrup, and Wasserstrom 2012). The increase in these rates is probably associated with hyperphosphorylation of RyRs by Ca^2+^/ calmodulin-dependent protein kinase II (CaMKII), whose activity is increased due to the greater [Ca^2+^] resulting from the altered location and functioning of NKA/NCX/ANK2 complex (Popescu et al. 2016) in the gap between the plasma membrane and SR. In addition, recently, ankirin-B has been described interacting with WNT/β-catenin pathway by modulating of β-catenin’s phosphorylation state: pharmacological activation of WNT/β-catenin pathway was able to rescue an animal model [Ank2-cKO] from arrhythmogenic cardiomyopathy (Roberts et al. 2019). β-catenin transcript was also identified as a hub-bottleneck in our main network, consistently upregulated in AF, adding evidence about the role of the deregulation of these proteins’ expression in the pathophysiology of AF.

Therefore, it is not surprising that studies evaluating the pharmacological efficacy of inhibiting these mechanisms in the context of heart disease show divergent results, suggesting that the protective and cytotoxic potential of these pathways are tenuously separated (Hedhli, Lizano, et al. 2008; Hedhli, Wang, et al. 2008; Tang et al. 2010; Carrier 2010). Linked to the induction of autophagic pathways is the protein damage resulting from oxidative stress. As in other cardiac pathologies, the balance between the production of reactive oxygen species (ROS) and the antioxidant capacity of the atria has been associated with the development and progression of AF (Korantzopoulos et al. 2018). A series of comorbidities commonly associated with AF lead to an increase in the production of ROS, such as hyperglycemia, dyslipidemia, hypertension, inflammation, and an increase in tissue levels of angiotensin II (Heijman et al. 2014; Korantzopoulos et al. 2018; Chiang et al. 2012). Oxidative stress predisposes atrial tissue to ectopic activity by altering the physiology of several ionic currents, such as Na^+^and K^+^, but mainly that of Ca^2 +^, affecting the activity of LTCCs and RyR, causing Ca^2+^leakage from RS and its subsequent arrhythmogenic effects (Bukowska et al. 2008; Xie et al. 2015; Anzai et al. 1998; Youn et al. 2013). It also contributes to the establishment of conduction disorders of AF due to exacerbation of cardiac fibrosis through the activation of myofibroblasts by TGF-*β* (Friedrichs, Baldus, and Klinke 2012). The pharmacological treatment of arrhythmic disorders via inhibition of oxidative stress, therefore, has been identified as promising (Korantzopoulos et al. 2018) and, in this meta-analysis, we point out the deregulated genes belonging to this biological pathway (Table S2), which configure potential study targets in the pathophysiology of AF. The deregulated *NOX4* gene for example, is upregulated in the oxidative stress response pathway identified in module 2 and encodes isoform 4 of the NADPH oxidase enzyme (Youn et al. 2013). Along with *NOX2*, *NOX4* is the most expressed isoform in cardiac tissue and is related to the induction of cardiac cell apoptosis and activation of myofibroblasts from the production of H_2_O_2_ (Youn et al. 2013; Zhang et al. 2012). Its high expression in AF, therefore, suggests the continuous oxidative injury to which the atrial tissue is subjected and also appears as a potential therapeutic target.

Enriching for several biological pathways in modules 2 and 3 is the gene *CTNNB1*, which codes for β-catenin. This gene is upregulated and is classified as a hub-bottleneck in the PPI network generated after meta-analysis. Wnt/β-catenin pathway regulates the cytosolic levels of β-catenin, which, in the absence of signal by Wnt binding proteins, is associated with the APC/CK-1/GSK-3 protein destruction complex that rapidly phosphorylates it, marking it for ubiquitination and, later, degradation. The activation of this pathway, in turn, disrupts the protein destruction complex, allowing the accumulation of β-catenin and its translocation to the nucleus, where it interacts and activates the TCF/Lef transcription factor, allowing its various target genes to be expressed (Barker 2008). Wnt/β-catenin pathway is evolutionarily conserved in animals and controls processes of cell differentiation, proliferation and survival and of tissue development and homeostasis. The genes and pathways it regulates, however, are tissue-dependent and modify according to the physiological context (Barker 2008; MacDonald, Tamai, and He 2009; Frietze et al. 2012). Since Wnt/β-catenin pathway is involved with the homeostasis of different tissues and since it regulates such fundamental processes, its deregulation is involved with several diseases and types of cancer (Nusse and Clevers 2017). At the heart, it plays a crucial role throughout embryonic development, but is quiescent in adults. However, stressful stimuli such as ischemic damage, oxidative stress and high pressure loads can reactivate it, contributing to the development of diseases such as hypertrophy, heart failure and arrhythmias (Barker 2008; Malekar et al. 2010; Dawson, Aflaki, and Nattel 2013). In the context of atrial fibrillation, a series of cellular changes induced by the Wnt/β-catenin pathway may be contributing to its establishment and progress. The increase in nuclear β-catenin levels worsened the oxidative damage induced in an animal model of diabetic cardiomyopathy by β-catenin/c-Myc interaction (Liu et al. 2017). Likewise in AF, the activation of this pathway may be contributing to the exacerbated oxidative damage. Recently, it was also observed that the transcription factor TCF/Lef activated by β-catenin negatively regulates the expression of the Nav1.5 sodium channel, encoded by the SCN5a gene, in addition to modulating its kinetics, promoting tachycardia and ventricular fibrillation in animal model (Huo et al. 2019). Regarding the structural remodeling of the atria, Wnt/β-catenin regulates the transcription of genes related to fibrosis in AF and in several other fibrotic diseases (Lv et al. 2019). There is evidence, therefore, to indicate that increased signaling by β-catenin may be a candidate for “chromatin remodeling” pathway enriched in module 3, since it regulates several processes and genes intrinsically linked to the pathophysiology of AF.

Under stress, the increased expression of β-catenin in the atria may be linked to a frustrated protective response through cell de-differentiation and subsequent transcription of genes involved in the proliferation, development and survival of cardiac tissue cells. This hypothesis is supported by our meta-analysis considering both the identification of differentially expressed genes and the enrichment of pathways in the network formed by them related to tissue development such as “cardiac muscle tissue development”; “cell-substrate adhesion”; “muscle cell differentiation”; “nuclear chromosome segregation”; “establishment of organelle localization”; “regulation of morphogenesis of an epithelium”; “positive regulation of neurogenesis”; “regulation of cell morphogenesis” and “regulation of morphogenesis of an epithelium”. The resistance of cardiac tissue to reactivation of embryonic and proliferative pathways, however, as well as the observed exacerbation of several pathological mechanisms, end up suggesting that it is the inhibition of signaling by β-catenin that should be studied aiming at the therapeutic scope of AF. Finally, the activation of proliferative pathways may also be explaining the high expression identified in this meta-analysis of the *CCNB1* gene, another hub-bottleneck in our network that codes for the regulatory protein of the cell cycle cyclin B1. In adult cardiomyocytes, the levels of this transcript are extremely low, since almost all cardiomyocytes are trapped in the G1/S or G2/M phases of the cell cycle (Ponnusamy, Li, and Wang 2017; Ahuja, Sdek, and MacLellan 2007). Forced expression of the cyclin B1/CDK1 complex promotes mitosis in vivo and in vitro models, but the ability to promote cytokinesis and, therefore, effective tissue regeneration seems limited (Bicknell, Coxon, and Brooks 2004). This gene’s topological importance in the network and its expression deregulation, however, can still be explored in the sense of representing an important biomarker in AF.

In the fourth module identified, enrichment of pathways related to leukocyte’s activation, regulation and migration is notable, shedding light on the chronic inflammatory activity occurring in AF. Several cardiac comorbidities associated with AF, such as hypertension, ischemic disease or atherosclerosis, as well as angiotensin II signaling, may play a role in promoting the systemic or atrial pro-inflammatory state (Andrade et al. 2014; da Silva 2017). Once the disease is established, the damage and tissue remodeling present in AF promotes inflammation through the leakage of “damage-associated molecular patterns” (DAMPs) (Hu et al. 2015). Cytopathological analyzes of atrial tissue in patients with AF reveal intense leukocyte infiltrate and necrosis of adjacent cardiomyocytes (Frustaci et al. 1997), which is congruent with the “cell-cell adhesion” regulation pathway enriched in module 4, which acts on the expression of adhesion proteins in lymphocytes. Once in the atrial tissue, these cells secrete high levels of TGF-β, which contributes to structural remodeling by activating myofibroblasts and IL-6, which induces electrical remodeling by decreasing the expression of connexins, for example (Jalloul and Refaat 2019). It is also known that *IL-6 also* induces the expression of prothrombotic proteins (Kerr, Stirling, and Ludlam 2001; Kaski and Arrebola-Moreno 2011). In this meta-analysis, the von Willebrand factor transcript (*VWF*) was identified upregulated and classified as hub-bottleneck, enriching in pathways related to its physiological function such as cell adhesion, homeostasis and platelet degranulation. Therefore, in addition to the slow and turbulent blood flow favoring the formation of thrombi and thromboembolic events in patients with AF, the high expression of *VWF* may contribute to this condition. In addition, it interacts in a central way with several other proteins in the PPI network generated from the AF differentially expressed genes, suggesting that the modulation of its levels may emerge as a therapeutic alternative involving pathways well beyond the classic ones related to the use of platelet aggregation inhibitors.

## CONCLUSIONS

In this meta-analysis, we integrated data from nine AF-related datasets obtained by microarray experiments to raise more robust hypotheses about genes and pathways involved in the pathophysiology of disease. Our analyses resulted in a list of 589 differentially expressed genes, which after PPI network investigation were associated with four functional modules and 11 hubs-bottlenecks genes with strong association to the pathophysiology of AF. The analysis of the biological processes enriched in these modules demonstrated good congruence with recent literature and shed light on key pathways and proteins in the context of the disease. Thus, our work corroborates the potential of meta-analysis of transcriptomic data in the study of the mechanisms that permeate complex diseases, and suggests news insights into the molecular basis of AF. We emphasize that a more in-depth analysis of our results, involving the experimental validation of selected findings, may drive significant advances regarding AF and therapeutic approaches.

## Data Availability

All datasets analyzed in the current study are publicly available in the Gene
Expression Omnibus (GEO) database

## Acknowledgements

The authors would like to thank the Brazilian funding agency CNPq for the financial support. RHB was a recipient of the CNPq/UFRGS Institutional Program of Initiation Scholarships in Technological Development and Innovation (PIBITI).

## Supplementary material

**Table S1.** List of all differentially expressed genes identified in the meta-analysis and their differential expression pattern (up- or down-regulation).

**Table S2.** Biological pathways enriched in each network module, their differentially expressed genes, and respective differential expression patterns.

**Figure S1.** Set size and gene intersections for the top 20 pathways enriched for a) Cluster 1, b) Cluster 2, c) Cluster 3, and d) Cluster 4.

## Declarations

### Data availability statement

All datasets analyzed in the current study are publicly available in the Gene Expression Omnibus (GEO) database.

### Authors’ contribution

RHB and MRM designed the experiment. RHB performed the experiment. RHB and MRM analyzed the data. RHB wrotethe paper. Both authors read and approved the manuscript.

### LIST OF ABBREVIATIONS

CaMKII: Ca2^+^/calmodulin-dependent protein kinase II
DADs: delayed afterdepolarizations
DAMPs: damage associated molecular patterns
AF: atrial fibrillation
HSPs: heat-shock proteins
LTCC: L-type Ca^2+^ channels
NCX: Na^+^/K^+^ pump
NKA: Na^+^/Ca^2+^ exchanger
ER: endoplasmic reticulum
ROS: reactive oxygen species
RS: sarcoplasmic reticulum
RyR: ryanodine receptor

